# Improved Type 2 Diabetes Risk Stratification in the Qatar Biobank Cohort by Ensemble Learning Classifier Incorporating Multi-Trait, Population-Specific, Polygenic Risk Scores

**DOI:** 10.1101/2023.06.23.23291830

**Authors:** Ikhlak Ahmed, Mubarak Ziab, Shahrad Taheri, Odette Chagoury, Sura A. Hussain, Jyothi Lakshmi, Ajaz A. Bhat, Khalid A. Fakhro, Ammira S. Al-Shabeeb Akil

## Abstract

**Background:** Type 2 Diabetes (T2D) is a pervasive chronic disease influenced by a complex interplay of environmental and genetic factors. To enhance T2D risk prediction, leveraging genetic information is essential, with polygenic risk scores (PRS) offering a promising tool for assessing individual genetic risk. Our study focuses on the comparison between multi-trait and single-trait PRS models and demonstrates how the incorporation of multi-trait PRS into risk prediction models can significantly augment T2D risk assessment accuracy and effectiveness.

**Methods:** We conducted genome-wide association studies (GWAS) on 12 distinct T2D-related traits within a cohort of 14,278 individuals, all sequenced under the Qatar Genome Programme (QGP). This in-depth genetic analysis yielded several novel genetic variants associated with T2D, which served as the foundation for constructing multiple weighted PRS models. To assess the cumulative risk from these predictors, we applied machine learning (ML) techniques, which allowed for a thorough risk assessment.

**Results:** Our research identified genetic variations tied to T2D risk and facilitated the construction of ML models integrating PRS predictors for an exhaustive risk evaluation. The top-performing ML model demonstrated a robust performance with an accuracy of 0.8549, AUC of 0.92, AUC-PR of 0.8522, and an F1 score of 0.757, reflecting its strong capacity to differentiate cases from controls. We are currently working on acquiring independent T2D cohorts to validate the efficacy of our final model.

**Conclusion:** Our research underscores the potential of PRS models in identifying individuals within the population who are at elevated risk of developing T2D and its associated complications. The use of multi-trait PRS and ML models for risk prediction could inform early interventions, potentially identifying T2D patients who stand to benefit most based on their individual genetic risk profile. This combined approach signifies a stride forward in the field of precision medicine, potentially enhancing T2D risk prediction, prevention, and management.

## INTRODUCTION

Type 2 Diabetes (T2D) is a global public health challenge due to its rapidly rising prevalence and related complications (Zimmet et al. 2014; El-Kebbi et al. 2021). There are many factors contributing to the incidence of T2D within the population, including lifestyle choices (Tuomilehto et al. 2001; Hu 2011), diet (Ley et al. 2014; Schwingshackl et al. 2017) and their interplay with genetic and epigenetic elements (Drong et al. 2012), demonstrating a critical role in the disease development (Upadhyay et al. 2018). The polygenic nature of T2D has been well-established, indicating that the disease is influenced by multiple genetic variants, each with a small effect size (Voight et al. 2010; Mahajan et al. 2018). Genome-wide association studies (GWAS) have played a significant role in unraveling the genetic architecture of T2D. These studies involve scanning the entire genome of individuals to identify common genetic variants associated with the disease (e.g., (Voight et al. 2010). Through GWAS, many single nucleotide polymorphisms (SNPs) that are significantly associated with T2D risk have been identified. These genetic variants are often located in or near genes involved in critical biological pathways related to insulin secretion, insulin resistance, and β-cell function (Mahajan et al., 2018; Voight et al., 2010).

Polygenic Risk Scores (PRS) are becoming increasingly relevant in the field of genomic medicine (Kumuthini et al. 2022). They summarize an individual’s risk for a particular disease based on their genome-wide genotype data and have been deployed with varying degrees of success for predicting the risk of numerous complex diseases, including type T2D (Torkamani et al. 2018; Khera et al. 2018). For instance, PRS has shown the ability to identify individuals with a three-fold increased risk for coronary artery disease and T2D (Khera et al. 2018). The construction of a multi-trait PRS for T2D demonstrated that this approach explained 18% of T2D heritability and outperformed the single-trait PRS (Mahajan et al. 2018). However, a considerable limitation of these studies lies in the predominance of populations of European descent in the datasets used to calculate PRS. Hence, the applicability of these scores to populations with diverse genetic architectures, such as those in the Middle East and North Africa (MENA) region, is questionable (Duncan et al., 2019; Martin et al., 2019).

There exists a significant gap in the literature regarding the application of risk models, including PRS, in MENA populations. The prevalence of T2D in these populations is alarmingly high, exceeding 12.2% in adults (El-Kebbi et al. 2021). This highlights the urgent need to develop population-specific risk prediction models that consider the unique genetic architecture of these populations. In particular, Qatar exhibits one of the highest global rates of T2D, with a prevalence of 22% (O’Beirne et al. 2016). The country is predominantly composed of three genetic subpopulations: Bedouin, Persian-South Asian, and African (Rodriguez-Flores et al. 2016; Hunter-Zinck et al. 2010). Qatar provides a compelling rationale for developing population-specific risk prediction models, particularly for T2D, as the limited contribution of European/Asian T2D SNPs to the high T2D prevalence in Qatar implies distinct genetic risks compared to Europeans and Asians (O’Beirne et al. 2016). This genetic diversity, coupled with the country’s rapid economic growth and lifestyle transitions, likely contributes to the heightened prevalence of T2D in Qatar.

To bridge this gap and address the specific challenges posed by the unique genomic landscape of Qatar, machine learning (ML) techniques, particularly ensemble learning, have emerged as a promising approach (Chen and Asch 2017). Artificial intelligence and ML algorithms have been integrated into data mining pipelines to develop predictive models for T2D complications, achieving an accuracy of up to 83.8% (Dagliati et al. 2018). Additionally, an improved feature space-based gradient boosting regression tree ensemble approach demonstrated an accuracy of 82.49% and better generalization ability compared to single models and other ensemble learning models, contributing to improved risk prediction of T2D complications (Wang et al., 2023). In addition, a novel algorithm utilizing an Entropy-based technique, ResNet architecture, and Support Vector Machine (SVM) achieved an accuracy rate of 99.09% in identifying T2D risk variants from spectrum images (Das 2022). Ensemble learning techniques combine multiple learning algorithms to achieve optimal predictive performance (Zhou 2012). These techniques have demonstrated a promising impact across various disease contexts, including T2D, improving disease prediction and outcome prognosis (Maniruzzaman et al. 2018). Therefore, these approaches could offer a robust method for multi-trait PRS development, accounting for the complex trait correlations in disease prediction models (Kotsiantis et al. 2006).

The (QGP), an ambitious initiative that aims to sequence the genomes of all Qatari citizens, provides an unprecedented resource for studying genetic factors contributing to T2D and other diseases within the context of a Middle Eastern population (Fakhro et al. 2016). By leveraging the wealth of genomic data from the QGP and considering the unique genomic landscape of Qatar, our study aims to utilize the power of ensemble learning to construct a classifier that integrates multi-trait PRS (Hunter-Zinck et al. 2010; O’Beirne et al. 2016). Utilizing the comprehensive genomic data from the Qatar Biobank cohort (QBB) and targeting the genomic variants linked to T2D, we aim to enhance T2D risk stratification in this unique population. The successful application and validation of the ensembl learning classifier in the context of this study will substantially improve T2D risk stratification. This endeavor has the potential to improve T2D risk prediction not only in Qatar but also in other similar populations within the MENA region. Subsequently, this could inform the development of effective preventative measures and enable the provision of targeted treatments, consequently improving health outcomes in this high-risk population.

## METHODS

### Cohort Description

This study was conducted on the latest release of the QBB (Al Thani et al. 2019) comprising more than 14k subjects, who were comprehensively phenotyped at QBB and their whole genomes sequenced through (QGP)(Mbarek et al. 2022). The QBB is a thorough longitudinal cohort study that focuses on a population sample of permanent inhabitants of Qatar. This sample comprises Qataris, Arabs from other Arab countries, and non-Arab residents with regular follow-up assessments conducted at five-year intervals. The QBB further includes sociodemographic information, clinical and behavioral phenotypic data, biological samples (such as blood, urine, saliva, DNA, RNA, viable cells, and others), clinical biomarkers, and Omics data (including genomics, transcriptomics, proteomics, metabolomics, and more) (Al Thani et al. 2019).

### Qatar Biobank Sample Collection

QBB collected both physical and clinical measurements, along with various biological samples. These biological samples consisted of approximately 60 ml of blood, 5 ml of saliva, and 10 ml of urine. Participants were instructed to fast for 8 hours before their visit, although due to different visit shifts, most of the samples were spot specimens. The blood samples were subjected to analysis in order to assess 66 different biomarkers associated with disease risk factors. The hematology and blood chemistry biomarkers were analyzed specifically at the laboratories of Hamad General Hospital. The blood samples collected using EDTA tubes were centrifuged to separate them into plasma, buffy coat (leukocytes), and erythrocytes. After collection, all the samples were aliquoted and stored in three different locations (Al Thani et al. 2019).

### Whole Genome Sequencing and Processing

At the Sidra Clinical Genomics Laboratory Sequencing Facility, the library construction and sequencing processes were carried out. After extracting genomic DNA (gDNA), the integrity of the samples was assessed using the gDNA assay on the Perkin Elmer Caliper Labchip GXII. The concentration of the DNA was measured using the Invitrogen Quant-iT dsDNA Assay on the FlexStation 3. For library construction, approximately 150 ng of DNA was utilized with the Illumina TruSeq DNA Nano kit. Each library was indexed using the Illumina TruSeq Single Indexes. The quality and concentration of the libraries were evaluated using the DNA 1k assay on a Perkin Elmer GX2. Quantification of the libraries was performed using the KAPA HiFi Library quantification kit on a Roche LightCycler 480 (Mbarek et al. 2022).

### Phenotype Data Processing

The phenotype data in the form of lab results and measurement data as well as accompanied subject and nurse questionnaire were obtained from QBB. As a first step, the data files obtained were merged to produce a single data frame to be queried for relevant phenotypes and integration with genotype data. We identified a set of 11 phenotypes as clinical risk factors for type 2 diabetes. These were BMI, waist-to-hip ratio (WHR), c-peptide, insulin, glucose, HbA1c, high-density lipoprotein cholesterol (HDL), low-density lipoprotein cholesterol (LDL), total testosterone, thyroid stimulating hormone (TSH) and triglycerides (TGL). The distribution of the raw phenotypes was non-normal and missing data was observed for all eleven traits. The phenotypic data was imputed using the R mice package (van Buuren and Groothuis-Oudshoorn 2011), which offers a variety of imputation approaches for multivariate data, to impute the missing data. It can impute mixtures of two-level, continuous, binary, ordered, and unordered categorical data. The mice function returns “m” imputed datasets after computing multiple imputations using chained equations on an incomplete dataset. We used 250 number of multiple imputations (m) and aggregated all the completed datasets returned by the function using the mean of the imputations to get a simple imputation. Finally, the rank-based Inverse Normal Transformation (INT) was applied on all quantitative phenotypes to obtain properly normalized distributions.

### Genotype Data Processing and Quality Control

Multiple quality control steps were performed using plink (Purcell et al. 2007) to identify a set of reliable markers and subjects to be used for final association testing. We started with ∼129 million short variants in 14669 individuals and after filtering for SNPs with greater than 10% missing per marker (plink --geno), we retained 124 million markers. We extracted autosomal SNPs and removed variants with low minor allele frequencies (MAF; (plink --maf 0.05 filter), these filters resulted in a net of 6,609,963 SNPs. Finally, Hardy-Weinberg equilibrium (HWE) check was also performed for both case and control subjects separately by applying two different stringency levels. We used an HWE cut-off of 1e-06 for controls while for cases a cut-off of 1e-10 was used. This resulted in a final set of 5,069,660 SNPs that were considered for the association analysis.

On subject level checks, we performed missingness per individual (plink --mind), followed by sex check as a quality control measure to identify any individuals with discrepancy between the reported gender and the expected gender as based on the X-chromosome heterozygosity/homozygosity rates. This helps to identify any sample mix-ups. Males should have an X chromosome homozygosity estimate >0.8 and females should have a value <0.2. Using this approach, we identified only few samples that had a mismatch between the reported and estimated gender, these samples were discarded for further analysis. Similarly, heterozygosity of individuals is an important measure that can indicate sample contamination or inbreeding. We plotted the distribution of heterozygosity rate in our subjects and removed individuals with a heterozygosity more than three standard deviation (±3 SD) from the mean. Checks for heterozygosity are performed on a set of SNPs which are not highly correlated. Therefore, to generate a list of non-(highly) correlated SNPs, high LD regions were first excluded. The remaining SNPs were pruned using 5 markers at a time in a window size of 50 and 0.2 as the multiple correlation coefficient of an SNP being regressed on all other SNPs. The output of the heterozygosity checks resulted in 112 individuals deviating more than 3 SD’s from the mean and comprised of 31 T2D and 81 normal subjects. After applying all the above filters, we were left with 14278 individuals.

### GWAS Analysis

The GWAS analysis was conducted on quality control processed genotype and phenotype data sets using the linear mixed-effect model (LMM) implemented in the BOLT-LMM version 2.3 package (Loh et al. 2015, 2018). For GWAS analysis of all traits except for the BMI and WHR, the covariates used in the associating test model were, age, gender, BMI, WHR, and the first ten principal components of the population. For BMI and WHR, only age, gender, and the first ten principal components were used as covariates.

## RESULTS

### Genome-Wide Association Analysis

This study used 14,278 individuals from the QBB cohort corresponding to 7968 females, 6310 males, and complied of 4455 diabetic/pre-diabetic cases (T2D) and 9823 controls. Individuals were classified as diabetic or prediabetic if they self-reported being clinically diagnosed or had hemoglobin A1c (HbA1c) levels equal to or more than 6.5%. To identify genomic variants associated with the disease risk, we conducted GWAS with the diabetes status and 11 other established risk factors for T2D, which included: BMI, waist-to-hip ratio (WHR), c-peptide, insulin, glucose, HbA1c, (HDL), (LDL), total testosterone, (TSH) and triglycerides. We first investigated the heritability estimates of these traits (*h^2^*) – the proportion of phenotypic variation explained by genetic variants using the GREML analysis implemented in GCTA (Yang et al. 2011) (Table 1). These heritability estimates ranged from 17% for insulin to 41% for HDL. Following this, the GWAS analysis for all 12 traits was conducted using LMM for ∼5 million common variants, while correcting for Age, sex, BMI, and population structure using first 10 principal components. Quantile-quantile (QQ) plots did not indicate any inflation in our results and genomic inflation factor (λ_GC_) ranged from 0.99 to 1.09 (Table 1). For all 12 traits, we identified 397 genomic loci that were in the vicinity of protein coding genes (within 1Mb) and showed association at genome-wide significance with at least one of the traits. For example, for the BMI GWAS we identified three top associated variants for genes MC4R (rs12964203; pvalue = 4.3E-10; beta = 0.08), TMEM18 (rs7570232; pvalue = 6.6E-09; beta= −0.08) and FTO (rs8050136; pvalue = 1.6E-08; beta= 0.064). Figure 1 shows the Manhattan plot for the BMI GWAS, with 86 variants associating with the trait at a genome-wide significance cut-off (5e-08). These 86 variants showed multiple independent hits in three genes Fat mass and obesity associated (*FTO*), Melanocortin 4 Receptor (MC4R) and Transmembrane Protein 18 (TMEM18) (Figure 1). Similarly, for T2D status we observed significant hits in two protein coding genes, for WHR three genes were identified, for HbA1c nine genes were observed (Table 2), and the number of associated genes rose significantly for other traits (Figure 1C).

**Figure 1:**
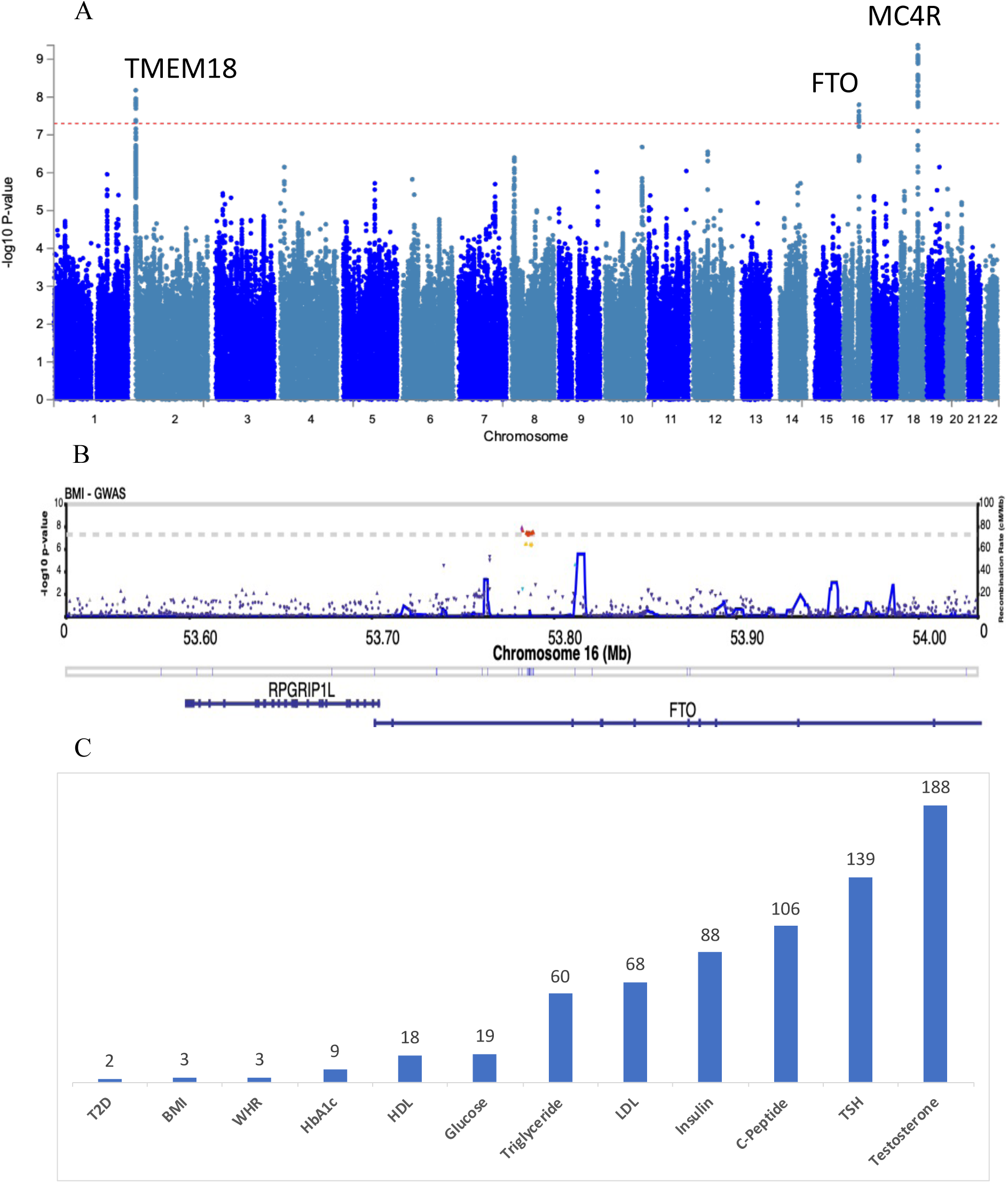
(A) Manhattan plot of a large-scale BMI GWAS. The plot depicts the negative log-base-10 of the P value for each polymorphism in the genome (along the x-axis) on the y-axis. The dashed line represents the significance level for genome-wide significance (P < 5 × 10^−8^). (B) Region association plot for the FTO gene. This plot shows a detailed view of the significant variants within FTO gene. The significant associations appear higher on the plot. Estimated recombination rate is shown in blue (right y-axis). Genomic position is shown on the x-axis. Red: Represents SNPs that have reached genome-wide statistical significance. Yellow: Indicates SNPs that approach statistical significance but did not reach the stringent thresholds. (C) Number of protein coding genes within 1Mb from genome-wide significant SNPs.

**Table 1.** Estimates for heritability of the traits and genomic inflation for GWAS.

|  |  |  |  |
| --- | --- | --- | --- |
| Trait | $h^2$ | SE | $\lambda_{GC}$ |
| T2D | 0.22 | 0.017 | 1.05 |
| BMI | 0.35 | 0.016 | 1.09 |
| C-peptide | 0.18 | 0.015 | 1.05 |
| Glucose | 0.22 | 0.015 | 1.05 |
| HbA1c | 0.36 | 0.016 | 1.09 |
| HDL | 0.41 | 0.017 | 1.09 |
| Insulin | 0.17 | 0.015 | 1.05 |
| LDL | 0.24 | 0.016 | 1.05 |
| Total testosterone | 0.20 | 0.015 | 1.00 |
| TSH | 0.31 | 0.016 | 1.09 |
| Triglycerides | 0.31 | 0.016 | 1.09 |
| WHR | 0.29 | 0.016 | 1.09 |

**Table 2.**
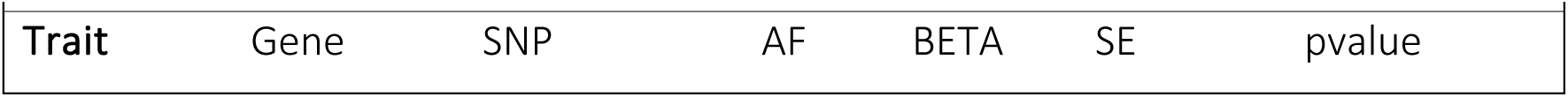
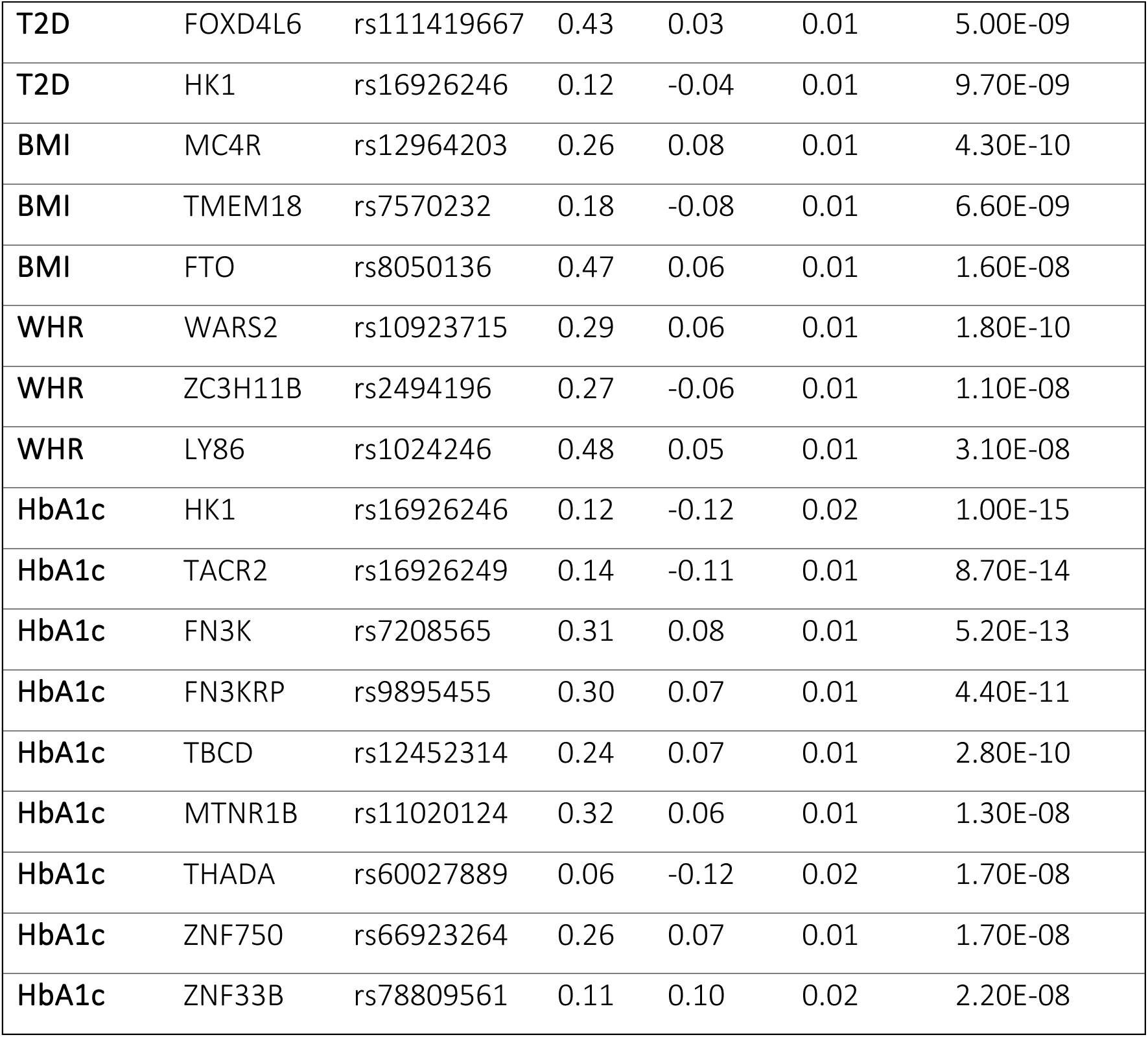
Top variants for each gene associating with the trait.

| Table 2. Top variants for each gene associating with the trait |  |  |  |  |  |  |
| --- | --- | --- | --- | --- | --- | --- |
| Trait | Gene | SNP | AF | BETA | SE | pvalue |
| **T2D** | FOXD4L6 | rs111419667 | 0.43 | 0.03 | 0.01 | 5.00E-09 |
| **T2D** | HK1 | rs16926246 | 0.12 | -0.04 | 0.01 | 9.70E-09 |
| **BMI** | MC4R | rs12964203 | 0.26 | 0.08 | 0.01 | 4.30E-10 |
| **BMI** | TMEM18 | rs7570232 | 0.18 | -0.08 | 0.01 | 6.60E-09 |
| **BMI** | FTO | rs8050136 | 0.47 | 0.06 | 0.01 | 1.60E-08 |
| **WHR** | WARS2 | rs10923715 | 0.29 | 0.06 | 0.01 | 1.80E-10 |
| **WHR** | ZC3H11B | rs2494196 | 0.27 | -0.06 | 0.01 | 1.10E-08 |
| **WHR** | LY86 | rs1024246 | 0.48 | 0.05 | 0.01 | 3.10E-08 |
| **HbA1c** | HK1 | rs16926246 | 0.12 | -0.12 | 0.02 | 1.00E-15 |
| **HbA1c** | TACR2 | rs16926249 | 0.14 | -0.11 | 0.01 | 8.70E-14 |
| **HbA1c** | FN3K | rs7208565 | 0.31 | 0.08 | 0.01 | 5.20E-13 |
| **HbA1c** | FN3KRP | rs9895455 | 0.30 | 0.07 | 0.01 | 4.40E-11 |
| **HbA1c** | TBCD | rs12452314 | 0.24 | 0.07 | 0.01 | 2.80E-10 |
| **HbA1c** | MTNR1B | rs11020124 | 0.32 | 0.06 | 0.01 | 1.30E-08 |
| **HbA1c** | THADA | rs60027889 | 0.06 | -0.12 | 0.02 | 1.70E-08 |
| **HbA1c** | ZNF750 | rs66923264 | 0.26 | 0.07 | 0.01 | 1.70E-08 |
| **HbA1c** | ZNF33B | rs78809561 | 0.11 | 0.10 | 0.02 | 2.20E-08 |

### Development of Polygenic Risk Scores

We created PRS for our diabetes cohort consisting of 4445 T2D subjects and 9823 controls based on GWAS summary stats for each of the 12 traits. PRS is an individual-level score that summarizes the effects of multiple genomic variants on the phenotype of an individual. Hence, the score is an estimate of the total genetic risk of a particular individual for a specific trait and thus can be used as an outcome predictor in clinical prediction and disease screening programs. We used PRSice 2(Choi and O’Reilly 2019) to identify the most predictive PRS based on the GWAS results of each trait. PRSice 2 is designed to calculate and evaluate the genetic risk scores based on GWAS summary statistics and individual-level genotype data. This involves clumping SNPs and performing P-value thresholding for thinning SNPs according to linkage disequilibrium and P-value. The clumping parameters --clump-kb 250kb and --clump-r2 0.1 were used to remove SNPs that are in linkage disequilibrium with one other, based on maximum likelihood haplotype frequency estimations (r^2^ values). To acquire the best PRS models from the GWAS results, multiple P-value thresholds were tried in order to identify the most optimal set of SNPs for the stratification of T2D cases and controls. We found that the best PRS models for each trait yielded results of 153970 SNPs for T2D, 158664 SNPs for HbA1c, 154054 SNPs for glucose, 104424 SNPs for triglycerides, 85672 SNPs for LDL, 155730 SNPs for insulin, 14112 SNPs for c-peptide, 127263 SNPs for TSH, 137767 SNPs for testosterone, 109488 SNPs for HDL, 9822 SNPs for WHR, and 78535 SNPs for BMI. Our PRS models displayed a high degree of stratification between cases and controls (Figure 2 A & B). The predictive power of the models is shown in (Figure 2C) and highlights the power gain by the PRS over the null model (using covariates Age, gender, BMI, WHR, first 10 PCs) at an estimated T2D prevalence (Awad et al. 2019) of 0.17 in our population. Empirical P-values obtained using 10,000 permutations of randomly shuffling phenotypic labels and repeating the analysis were used to control for Type 1 error. The empirical association *P-*values of all PRS models were 9.9e-05, the minimum possible, and hence did not show any indications of inflation or overfitting.

**Figure 2:**
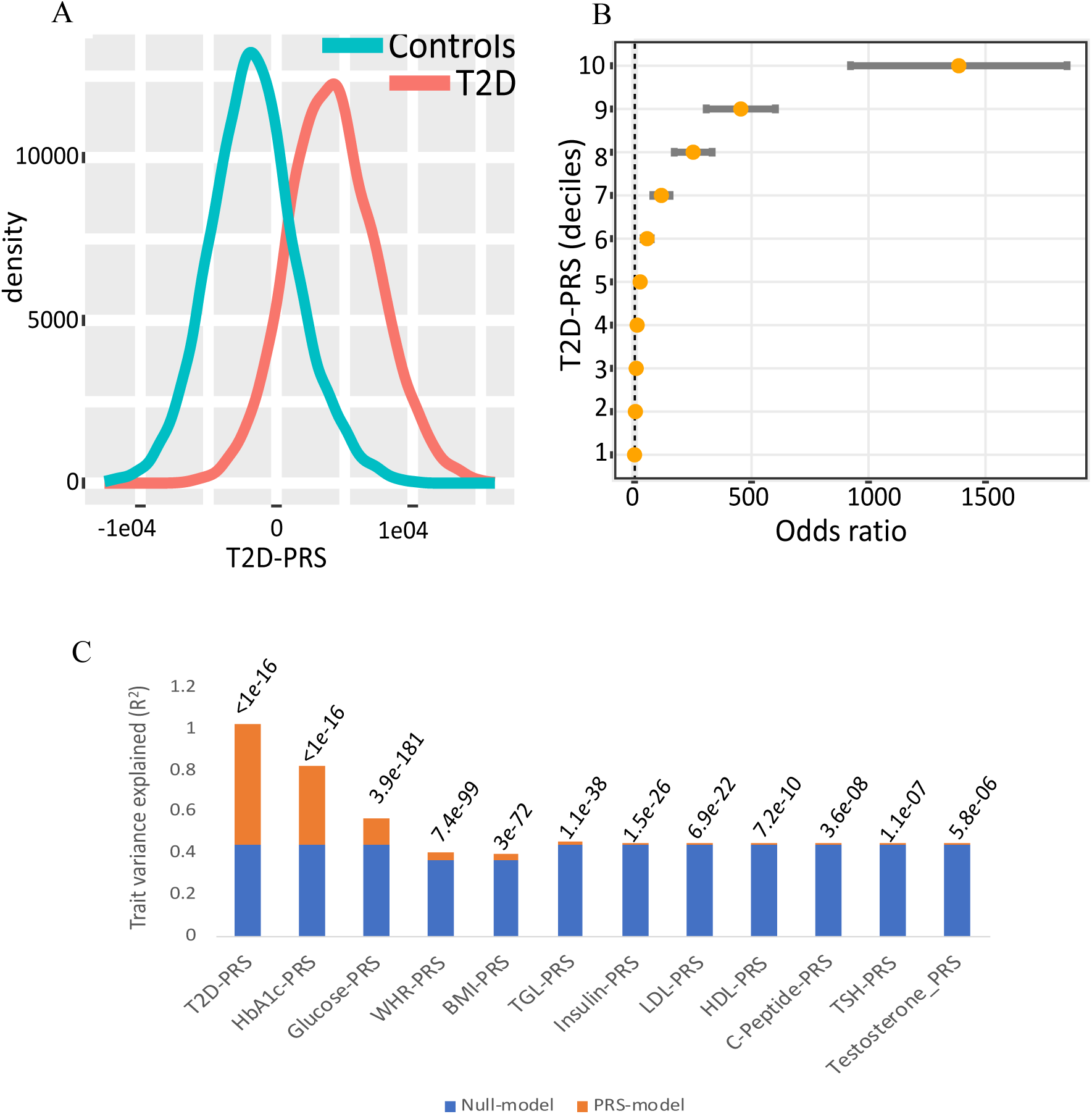
(A) Density plot showing distribution of T2D PRS scores in the QBB cohort stratified for cases and controls. (B) Strata plot. The Y-axis shows deciles (e.g., decile 9 corresponds to those individuals with PRS between the 90th and 100th percentile of the population), and the X-axis shows the odds ratio as obtained from the regression model Phenotype ∼ decile + covariates (C) Trait variance explained by PRS models. The stacked bar shows variance explained (R^2^) by the null model and the PRS model for each trait. Null model is the variance explained by covariates only, while the PRS-model is defined as the R^2^ of the Full model minus the R^2^ of the Null model. P-values in the plot indicate p values of the model fit.

**Figure 3.**
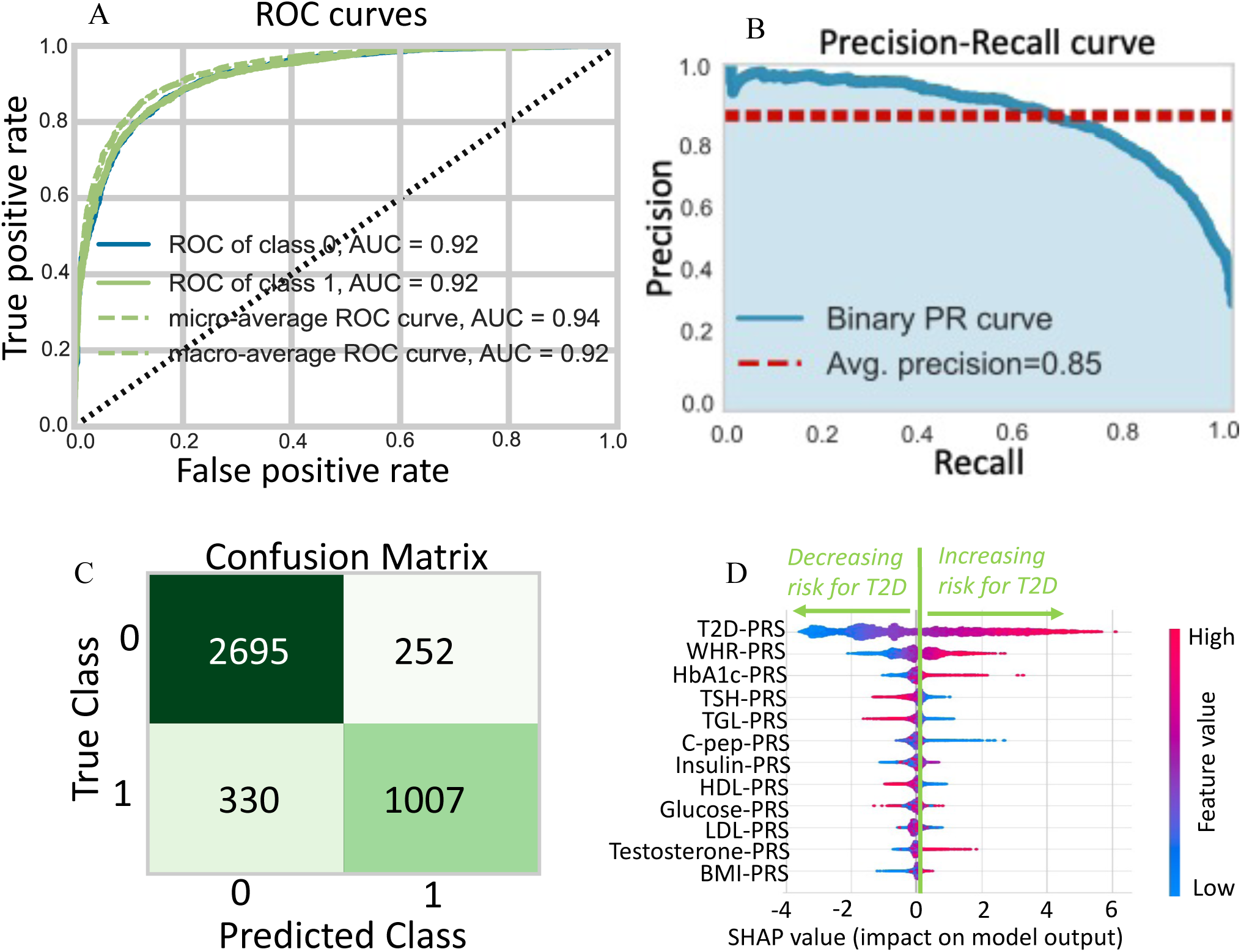
(A) The CatBoost classifier’s performance is depicted by the Receiver Operating Characteristic (ROC) curve. It depicts two values at different thresholds: True Positive Rate (TPR) (on the Y-axis) and False Positive Rate (FPR) (on the X-axis). TPR is proportion of observations that were correctly predicted to be positive out of all positive observations (TP/(TP+FN)), FPR is the proportion of observations that were incorrectly predicted to be positive out of all negative observations (FP/(FP+TN)). The curve is generated by plotting the TPR versus the FPR at various threshold values. The area under the ROC curve (AUC) indicates how well the model can differentiate between two classes. (B) Precision-Recall curve (PRC) evaluating the effectiveness of the CatBoost classifier. Precision shows the model’s accuracy in making positive predictions. A high accuracy score means that the model produces fewer false positives. Recall represents the model’s ability to correctly detect positive instances. A high recall score indicates that the model produces fewer false negatives. A higher AUC-PR indicates better model performance. (C) Confusion Matrix. The table shows, how predicted values by the model compare against the actual values for the test data. (D) SHAP plot: Bee-swarm plot of the SHAP (SHAPley additive explanations) values that shows the contribution of each PRS to the model’s predictive performance. Each dot for each feature represents a single individual and the width corresponds to the number of individuals with a given SHAP value. The color of the dot reflects the feature value relative to the median value of the feature, while the position on the x-axis indicates SHAP value. Positive SHAP values indicate an increased risk for T2D while negative values are associated with a lower risk for the disease.

### Multi-PRS Using Machine Learning

While PRS based on a single trait can be informative, it may not capture the full complexity of disease risk. To address this issue, we went one step further and created a multi-trait PRS model utilizing machine learning techniques to better stratify risk. We created sixteen different machine learning models and evaluated the performance of each model using a 10-fold stratified cross-validation at a train-test split of 70:30. Of the 16 models tested, CatBoost classifier was the best performer, followed by Random Forest, LightGBM, and XGBoost. Each model included 12 polygenic risk score (PRS) features. The best model achieved impressive results, with an accuracy of 0.8549, an area under a receiver operating characteristic curve (AUC) of 0.92, an area under the precision-recall curve (AUC-PR) of 0.8522, and an F1 score of 0.757 at a precision of 0.79 and recall of 0.73. The top features used in this model included PRS scores for T2D status, waist-hip ratio (WHR) and HbA1c. Surprisingly, the PRS for BMI was not one of the top features. This suggests that those who are genetically predisposed to having a high BMI may not be at an increased risk of developing T2D in our population. This is an intriguing result and could have implications for how we approach preventing the onset of T2D in the future.

## DISCUSSION

T2D is a multifaceted and complex metabolic condition that affects millions of people throughout the world. While environmental variables such as nutrition and lifestyle play an important role in its development, it is becoming increasingly evident that genetics has an equally important role in its susceptibility (Vassy et al. 2014; Stančáková and Laakso 2016; Udler 2019). GWAS has emerged as a powerful tool to identify genetic variants associated with T2D and its complications and has aided in understanding the underlying biology for the disease, improving risk prediction, and advancing personalized medicine approaches (Wheeler and Barroso 2011; Stančáková and Laakso 2016; Scott et al. 2017; Chen et al. 2019). Multiple GWAS studies have examined the genomes of thousands of individuals, and specific genetic variants that are statistically linked to T2D were identified. These findings have revealed important insights into the biological pathways involved in T2D development, including beta-cell dysfunction, insulin resistance, and impaired glucose metabolism [(Fuchsberger et al. 2016; Scott et al. 2017; Mahajan et al. 2018).

In this study, we conducted twelve robust GWASes to identify genetic loci that can impact the risk of T2D in our population. To achieve this, we conducted these studies on common genetic variants that included 11 risk factors and the T2D status. Prior to GWAS, stringent quality control measures were taken to ensure accuracy and reliability in the results. We chose to use common variants, as low MAF SNPs are both rare and more likely to have genotyping errors. In addition, HWE checks were conducted for both case and control subjects separately, using two different levels of stringency. Deviations from HWE can signify either genotyping errors or true association signals, so for control subjects, a stringent value of HWE (less than 1e-6) was used while case cohort was filtered for a less stringent HWE threshold (1e-10). This less stringent threshold for cases helps to avoid discarding true disease-associated SNPs under selection. We also conducted subject level checks for missingness, sex discrepancy and heterozygosity. Average heterozygosity of our cohort was observed to be 0.284 with a standard deviation of 0.0156. This helped identify 112 outliers that were removed from the subsequent analysis. For the 112 samples removed, 56% were with high heterozygosity indicating low sample quality while 44% had low heterozygosity indicating inbreeding. Any investigation of quantitative trait loci, such as GWAS, relies on the assumption that the phenotype data is distributed normally. If this assumption is violated, the power and type I error of the analysis can be significantly compromised. Additionally, the sample size available for the trait of interest will affect the power of the GWAS, as missing phenotypic data can lead to a lack of statistical power. In our data, the missingness rates ranged from 0.15 percent for BMI to 1.16 percent for HbA1c, and, although low, we still chose to impute the missing values to retain the entire dataset. The imputed values showed no signs of skewness across multiple imputations.

PRS have emerged as a powerful tool in genetics research, enabling the aggregation of genetic information to estimate an individual’s genetic susceptibility to specific traits or diseases. While traditional PRS primarily focus on single traits or diseases, there has been a growing interest in the development of multi-trait PRS. This advancement holds great potential in enhancing our understanding of complex traits, improving risk prediction, and identifying shared genetic mechanisms. The multi-trait PRS can help identify novel links between risk factors and T2D as well as enable the identification of shared genetic variants and pathways that contribute to multiple traits or risk factors for a disease. By considering the shared genetic architecture across different traits and risk factors, previously unrecognized connections and associations could be uncovered. This broader perspective allows for a more comprehensive understanding of the complex interactions between genetic variants, risk factors, and the development of T2D. Thus, this approach not only enhances our understanding of the genetic underpinnings of T2D but also holds the potential to unravel the interconnectedness of biological processes involved in the disease.

By utilizing 12 different GWASes, we have identified sets of genetic loci that could influence the risk for developing T2D. These clusters of genetic loci not only exhibit associations with T2D susceptibility and its risk factors but could collectively contribute to the overall risk profile of an individual for T2D. We used multiple P-value thresholds to determine the best-fit PRS for each trait, resulting in maximum segregation of T2D cases and controls. The PRS models displayed a significant predictive power over the null model with the minimum possible empirical association P-value of 9.9e-05 from 10,000 permutations, which indicates that there was no inflation or overfitting in our models due to type1 error. These PRS models were further integrated into a multi-trait PRS using Machine learning approaches. The training and testing for each ML model was performed on a 70:30 split of the data. After conducting an extensive comparison of 16 different ML models, we identified CatBoost Classifier as the best performing model for our dataset. The model showed an accuracy of 0.8549, to correctly classify 85.49% of the samples, demonstrated its good ability to distinguish between positive and negative classes (AUC = 0.92). Moreover, the model also had a good balance between precision and recall (AUC-PR = 0.8522, F1 = 0.757), suggesting its potential to provide effective predictions in real-world applications. Our final model trained on the entire QBB dataset has shown even better performance, boasting an AUC of 0.97. As these results are significantly impressive, multiple validations are planned to comprehensively assess its performance on unseen data and ensure the reliability and generalizability of the model. We are now in the process of collecting independent T2D cohorts in order to validate the efficacy of the model and ensure that it is not overfitting. Once these validations are completed, we can be assured of the model’s performance and start utilizing it to gain insights and further our understanding of type 2 diabetes.

The results of our analysis were also quite surprising in terms of the feature importance for the model, as the top features used in the model to predict the T2D risk did not include the PRS for BMI. This suggests that those who are genetically predisposed to having a high BMI may not be as susceptible to developing T2D in our population. This is a fascinating finding as it could have implications for how we approach the prevention of T2D in the future. Instead of focusing solely on BMI, genetic predisposition for factors such as WHR and HbA1c, might be better predictors for assessing T2D risk in our population. Therefore, it is important to take into account the genetic variants associated with multiple risk factors of T2D to increase our understanding of this condition and to develop better prevention strategies.

## CONCLUSIONS

To improve the predictability of T2D risk, our study illustrates the significant potential and power of combining multi-trait PRS with ML. In contrast to single-trait PRS models, multiple trait-based PRS models have a significant advantage in finding genetic variants linked to T2D. The predictive power of our top ML model was evidenced by the impressive metrics, including an accuracy of 0.8549, an AUC of 0.92, an AUC-PR of 0.8522, and an F1 score of 0.757. These values are indicative of the model’s superior performance in stratifying cases from controls, providing a robust tool for T2D risk assessment.

These findings represent a significant stride forward in the field of precision medicine for T2D. By providing a more comprehensive, accurate, and individualized risk assessment, such models can identify high-risk individuals who stand to gain the most from early interventions. Thus, this research signals a paradigm shift in T2D risk management from a “one-size-fits-all” approach to one that is more individualized and targeted.

However, as with all predictive models, external validation is crucial for assessing their generalizability. Our ongoing efforts aim to validate the final model’s performance using independent T2D cohorts. Success in this endeavor will solidify our model’s utility and robustness, further cementing the role of multi-trait PRS and ML in T2D risk prediction.

The emergence of multi-trait PRS in genetics research has brought about exciting possibilities. By considering the shared genetic architecture across multiple traits or risk factors, multi-trait PRS offers a more comprehensive understanding of complex traits such as T2D. This approach enables the identification of shared genetic variants and pathways, fostering the discovery of novel links between risk factors and the disease. Ultimately, multi-trait PRS models could be instrumental in informing preventive strategies, fostering early interventions and personalized approaches to its prevention and treatment, and ultimately reducing the societal and health burden of T2D.

## AVAILABILITY OF DATA AND MATERIALS

Data described in the manuscript, including all relevant raw data, will be freely available to any scientist wishing to use them for non-commercial purposes without breaching participant confidentiality. Requests should be sent directly to the corresponding author.

## ABBREVIATIONS

T2D: Type 2 Diabetes
PRS: polygenic risk scores
GWAS: genome-wide association studies
QGP: Qatar Genome Programme
ML: machine learning
AUC: Area under the ROC Curve
SNPs: nucleotide polymorphisms
MENA: Middle East and North Africa
SVM: Support Vector Machine
QBB: Qatar Biobank cohort
HMC: Hamad Medical Corporation
WGS: Whole genome sequencing
HDL: high-density lipoprotein cholesterol
LDL: low-density lipoprotein cholesterol
TSH: Thyroid stimulating hormone
TGL: Triglycerides
INT: Inverse Normal Transformation
HWE: Hardy-Weinberg equilibrium
LMM: linear mixed-effect model
FTO: Fat mass and obesity associated
*MC4R*: Melanocortin 4 Receptor
TMEM18: Transmembrane Protein 18
WHR: Waist-Hip Ratio

## ACKNOWLEDGMENT

The authors would like to thank and acknowledge the support of the Qatar National Research Fund (QNRF) and Qatar Genome Program for funding the study under the grant PPM 03-0311-190017. We also would like to thank Qatar Biobank, which provided clinical and phenotypic data used in this study. The authors would like to thank Sidra Medicine, for providing the infrastructure that facilitated completing this work (Grant number SDR1000043, *The QGPRS Study; Qatar Genome Polygenic Risk Score: A Precision Medicine Approach to Prevent Diabetic Complications in the Affected Qatari Individuals*).

## FUNDING

This research was funded by the Path Towards Precision Medicine (PPM) of Qatar National Research Fund (QNRF), a subsidiary of Qatar Research Development and Innovation Council (QRDI) and Sidra Medicine Research Division, grants number PPM 03-0311-190017, and SDR1000043, respectively. Author A.S.A is the project lead principal investigator, budget holder, and the recipient of the research funding support from both organizations.

## AUTHOR CONTRIBUTION

Ikhlak Ahmed, manuscript writing and conducting the bioinformatic data analysis. Mubarak Ziab, contributed to the manuscript writing and visualization. Hamdi Mbarak, managing the genomic data, facilitating collaborations, and preparing the validation cohort for ongoing studies. Shahrad Taheri, forming the clinical validation cohort and selecting participants. Odette Chagoury managed the validation cohort and collected samples. Sura A. Hussain and Jyothi Lakshmi provided clinical research coordination at the main study site and collected data. Ajaz A. Bhat, contributed to manuscript writing, reviewing, and editing. Khalid A. Fakhro, contributed to the main study concept and reviewed and edited the manuscript. *Ammira S. Al-Shabeeb Akil, the conceptualization of the study, designing the cohort, designing the original study, acquiring funding, contributing to manuscript writing, reviewing, and editing, supervising the study team, and preparing the cohorts for the validation studies. All authors have read and approved the final manuscript.

## CORRESPONDING AUTHOR

Correspondence to Ammira Al-Shabeeb Akil

## ETHICS APPROVAL AND CONSENT TO PARTICIPATE

Informed consent was obtained from all participants included in the study through Qatar Biobank-“QBB Cohort Study QF-QBB-RES-ACC-0075 QBB IRB Approval number is: Full Board-2017-QF-QBB-RES-ACC-0075-0023”. Deidentified research conducted under the project “E-2020-QF-QBB-RES-ACC-0150-0143. QBB IRB Approval number, E-2017-QF-QBB-RES-ACC-0026-0001”. Sidra IRB MOPH Assurance: IRB-A-Sidra-2019-0020 Sidra IRB MOPH Registration: IRB-Sidra-2020-009 Sidra IRB DHHS Assurance: FWA00022378. Sidra IRB DHHS Registration: IRB00009930. Tel: +974-4003-7747. Sidra Medicine, IRB protocol # 1660756, May 30, 2024. Approval category: Expedited.

## CONSENT FOR PUBLICATION

Not Applicable

## COMPETING INTERESTS

The authors declare no conflict of interest

## Notes

* This work is supported by the PPM award (PPM 03-0311-190017) from QNRF to Dr. Ammira Al-Shabeeb Akil

### Competing Interest Statement

The authors have declared no competing interest.

### Author Declarations

QBB Cohort Study QF-QBB-RES-ACC-0075 QBB IRB Approval number is: Full Board-2017-QF-QBB-RES-ACC-0075-0023. Deidentified research conducted under the project E-2020-QF-QBB-RES-ACC-0150-0143. QBB IRB Approval number, E-2017-QF-QBB-RES-ACC-0026-000. Sidra IRB MOPH Assurance: IRB-A-Sidra-2019-0020 Sidra IRB MOPH Registration: IRB-Sidra-2020-009 Sidra IRB DHHS Assurance: FWA00022378. Sidra IRB DHHS Registration: IRB00009930. Sidra Medicine, IRB protocol # 1660756, May 30, 2024. Approval category: Expedited.

### Summary of Updates

No changes except removing one author based on his request due to his involvement in other conflicting project.

